# Fat burning capacity in a mixed macronutrient meal protocol does not reflect metabolic flexibility in women who are overweight or obese

**DOI:** 10.1101/2024.08.29.24312791

**Authors:** Mary M. Ahern, Virginia M. Artegoitia, Rémy Bosviel, John W. Newman, Nancy L. Keim, Sridevi Krishnan

**Author notes:** Corresponding author information: Sridevi Krishnan. co-first authors (Mary M. Ahern, Virginia M. Artegoitia).

## Abstract

**Introduction:** Metabolic flexibility, the ability to switch from glucose to fat as a fuel source, is considered a marker of metabolic health. Higher fat oxidation is often associated with greater flexibility and insulin sensitivity, while lower fat oxidation is linked to metabolic inflexibility and insulin resistance. However, our study challenges the universal validity of this relationship, uncovering a more nuanced understanding of the complex interplay between fuel source switching and fat oxidation, especially in the presence of insulin resistance.

**Methods:** In an 8-week controlled feeding intervention, overweight to obese women with insulin resistance (as defined by McAuley’s index) were randomized to consume either a diet based on the Dietary Guidelines for Americans 2010 (DGA) or a ‘Typical’ American Diet (TAD), n = 22 each. Participants were given a high-fat mixed macronutrient challenge test (MMCT) (60% fat, 28% carbohydrates, and 12% protein) at weeks 0, 2, and 8. Plasma lipids, metabolome, and lipidome were measured at 0, 0.5, 3, and 6h postprandial (PP); substrate oxidation measures were also recorded at 0,1 3, and 6h PP. Metabolic flexibility was evaluated as the change in fat oxidation from fasting to PP. Mixed model and multivariate analyses were used to evaluate the effect of diet on these outcomes, and to identify variables of interest to metabolic flexibility.

**Results:** Intervention diets (DGA and TAD) did not differentially affect substrate oxidation or metabolic flexibility, and equivalence tests indicated that groups could be combined for subsequent analyses. Participants were classified into three groups based on the % of consumed MMCT fat was oxidized in the 6h post meal period at weeks 0, 2 and 8. Low fat burners (LB, n = 6, burned <30% of fat in MMCT) and high fat burners (HB, n = 7, burned > 40% of fat in MMCT) at all weeks. Compared to LB, HB group had higher fat mass, total mass, lean mass, BMI, lower HDLc and lower RER (p < 0.05), but not different % body fat or % lean mass. During week 0, at 1h PP, LB had an increase in % fat oxidation change from 0h compared to HB (p<0.05), suggesting higher metabolic flexibility. This difference disappeared later in the PP phase, and we did not detect this beyond week 0. Partial least squares discriminant analysis (PLSDA (regular and repeated measures (sPLSDA)) models identified that LB group, in the late PP phase, was associated with higher rates of disappearance of acylcarnitines (AC) and lysophosphatidylcholines (LPC) from plasma (Q2: 0.20, R^2^X: 0.177, R^2^Y: 0.716).

**Conclusion:** In women with insulin resistance, a high fat burning capacity does not imply high metabolic flexibility, and not all women with insulin resistance are metabolically inflexible. LPCs and ACs are promising biomarkers of metabolic flexibility.

## Introduction

Metabolic flexibility refers to the ability of an organism to adapt to the energetic push and pull and switching between fuel substrates allowing for an efficient metabolic response to physiological needs and opportunity^1^. This flexibility is an important component of metabolic health, being a key metabolic adaptation that balances choice of fuel storage and oxidation. It can be measured as the shift in metabolism to use the more abundant fuel present in a given meal, e.g. fat, rather than defaulting to glucose^2^.

The functional capacity of skeletal muscle tissue, the most metabolically active tissue, plays a key role in metabolic flexibility^3^. The muscle of lean individuals is considered metabolically flexible, as it readily adapts to available fuels in response to insulin^4^. In contrast, obese-insulin resistant individuals are metabolically inflexible, and experience stunted insulin-stimulated suppression of fat oxidation following a meal with glucose. Metabolic flexibility is also closely linked to fat oxidation rates. Lower rates of maximal fat oxidation are associated with reduced metabolic flexibility^5,6^. Additionally, impaired insulin sensitivity and metabolic flexibility are linked to reduced fat oxidation under resting conditions in skeletal muscle^6^. Collectively, this suggests that skeletal muscle flexibility and the ability to oxidize lipids effectively are inextricably related to metabolic flexibility and this relationship ultimately impacts an individual’s health.

Perturbations in metabolic flexibility are associated with insulin resistance, metabolic dysfunction-associated fatty liver disease (MAFLD), type 2 diabetes and cardiovascular disease ^7–12^. Accurate measurement of metabolic flexibility can allow for its use as a biomarker of health. Metabolic flexibility is measured systemically in humans using indirect calorimetry during a euglycemic-hyperinsulinemic clamp^13^. Oral glucose tolerance tests, fasting and re-feeding tests, and oxygen restriction tests have also been used^14^. Currently, however, there is no consensus on when and how to measure metabolic flexibility.

Diet, especially fat composition, may impact metabolic flexibility. In mice, a high saturated fatty acid (SFA)-rich diet compared to a polyunsaturated fatty acid (PUFA)-rich diet reduced metabolic flexibility while increasing adiposity, liver damage and visceral fat deposits^18^. However, in humans, high-fat diets appear to be more useful as metabolic tests for long-term health^17^ and whole diet approaches, without weight loss, have not been successful at altering metabolic flexibility^19^. High-fat metabolic stress tests may be an effective way of measuring metabolic flexibility due to the stimulation of many metabolic systems simultaneously. Researchers have studied the association between dietary or meal fatty acid composition and subsequent systemic substrate oxidation^12,20–22^ but only a few studies have looked closely at circulating fatty acids and substrate oxidation^23,24^. The rise of high throughput omic tools has made it possible to get molecular insight into metabolic states. Subsequently, recent studies have attempted to deconstruct metabolic flexibility using lipidomic and metabolomic tools especially after a high-fat meal challenge test^16,25,26^.

This manuscript presents secondary outcomes from a randomized clinical trial that has been published^27^. The clinical trial compared metabolic effects of following a diet based on the Dietary Guidelines for Americans (DGA) to a Typical American diet (TAD) for 8-weeks in women (n = 44, 22 each group) with insulin resistance, as defined by McAuley’s index ^28^. The provided DGA diet was notably different in fat (less total fat, less saturated, more mono- and poly-unsaturated with more omega-3’s) than the TAD diet^29^. Here, we present our observations of metabolic rate parameters including fat oxidation and metabolic flexibility (calculated as the difference between measured fasting and postprandial fat oxidation^30^) in response to high-fat mixed macronutrient meal challenge tests (MMCT) conducted thrice during this diet intervention. In addition, fasting and postprandial lipidomic and metabolomic parameters in response to the MMCT were also measured. Our aims were to (a) identify if there was any effect of the diet intervention on metabolic rate, substrate oxidation and metabolic flexibility in response to the MMCT, and (b) glean further metabolic insight into fat oxidation and metabolic flexibility in women with insulin resistance using omic measures. As our intervention was designed to maintain body weight and physical activity during the testing period, we anticipated that our dietary intervention would have no effect on metabolic flexibility, especially given our small sample size^31–33^. Instead, we explored the metabolic rate and substrate oxidation responses to the repeated MMCT, with molecular input from lipidomic and metabolomic measures. To our knowledge, this is the first report to include three repeats of the same MMCT across weeks, and with metabolomic and lipidomic measures to enable identification of “stable” substrate oxidation and metabolic flexibility characteristics in individuals. By leveraging these metabolomic and lipidomic measures, we hope to enhance our understanding of this population’s underlying metabolic response to a high fat challenge test, thereby identifying potential biomarkers or metabolic signatures for further exploration.

## Methods

### Study design and participants

To test the impact of diets meeting the Dietary Guidelines for Americans on cardiometabolic risk factors, the individual Metabolism and Physiological Signatures Study (iMAPS; ClinicalTrials.gov: NCT02298725) recruited women who were aged 20–65 y with BMIs between 25–39.9 kg/m^2^. In addition, physical activity was limited to < 150 min/week, and they were insulin resistant based on screening tests to calculate McAuley’s insulin sensitivity index (values > 5.8 was considered insulin resistant^34^) ^27^. Other inclusion criteria included resting blood pressure ≤ 140/90 mm Hg, impaired glucose homeostasis and/or elevated fasting TGs, maintenance of a sedentary lifestyle with activity monitoring for 7-day periods four times over the study period using waist-worn accelerometers (Respironics® Actical™; Philips North America Co, Cambridge MA). Body composition was determined at 0 and 8 weeks by dual-energy X-ray absorptiometry (DXA; Hologic Discovery QDR Series 84994; Hologic, Inc.). A consort diagram describing participants is shown in **Supplemental Figure 1**.

### Mixed macronutrient challenge test

A high fat mixed macronutrient challenge vehicle contained 840 Kcals with 60% energy (calories) from fat, 28% from carbohydrates and 12% from protein was used in the mixed macronutrient challenge test (MMCT). The MMCT protocol was administered at week 0 prior to intervention, at intervention week 2, and intervention week 8 as reported^35^. On the evening before the test day, participants consumed a provided standardized pretest dinner and began a 12h fast. The following morning, after obtaining a fasting blood sample and measuring resting metabolic rate, the MMCT ‘milkshake-like’ meal was provided, and participants were given 10 minutes to consume it. After weighing back the residual left in the container, participants consumed 56 ± 2 g of palm oil, 59 ± 2 g of sucrose and 26 ± 1 g of egg white protein. The fatty acid composition by weight of the total fat content was 43% palmitate (16:0), 40% oleate (C18:1n9), 9% linoleate (18:2n6), 4% stearate (18:0), and < 1 % other detected fatty acid residues. The protocol included collecting four blood samples at 0 (fasting), and 0.5, 3 and 6h post meal challenge.

### Indirect calorimetry and metabolic flexibility

Estimates of fuel utilization were generated based on indirect calorimetry measures using automated metabolic carts with an open circuit system (TrueOne 2400, ParvoMedics). Measurements were taken four times for intervals of 15-20 min; the time sequence was fasting 0h, consumption of challenge meal, then 0.75-1h, 3h, and 6h after the MMCT, closely coinciding with blood collection times. Respiratory exchange ratio (RER), a common indicator of carbohydrate vs. fat combustion, was calculated as the ratio of measured volume of carbon dioxide (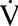CO2) produced to volume of oxygen (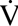O2) consumed using the equation 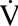CO2/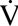O2. The resting and postprandial energy expenditure (EE) were estimated using the Weir equation without urinary nitrogen correction: EE = [(3.94 x 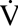O2) + (1.1 x 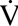CO2)]^36^. Rates of fat oxidation and carbohydrate oxidation were estimated using the Frayn equations^37^. A urinary nitrogen correction was used with these equations based on the protein content of the controlled diet, assuming participants were in nitrogen balance^29^. As mentioned earlier, metabolic flexibility was calculated as the change in postprandial fat oxidation compared to fasting, in response to the MMCT (i.e. postprandial – fasting fat oxidation)^30^. In addition, we also calculated % change in fat oxidation postprandial compared to fasting and change and % change in RER postprandial compared to fasting at all weeks.

### Plasma fatty acid analysis

Fatty acids were isolated in the presence of internal standards and quantified by gas chromatography-mass spectrometry as fatty acid methyl esters (FAMEs) against authentic calibration standards. Samples were processed in a total of 12 batches, each containing blanks, replicates, and laboratory reference materials. Samples were prepared using standard extraction and derivatization methods which are explained in greater detail in the supplemental methods.

Except for the non-esterified fatty acids (NEFA), surrogate recoveries, replicate precision and blank levels were acceptable. For NEFA, a subset of saturated fatty acids were compromised and excluded from the analysis. Subtle batch specific differences in NEFA were removed by adjusting samples’ means by laboratory reference material batch averages.

### Kit based-targeted metabolomics

Plasma concentrations of acyl carnitines (n = 40), amino acids (n = 21), biogenic amines (n = 21), glycerophosphospholipids (n = 90), sphingomeylins (n = 15) and total hexoses were measured using AbsoluteIDQ® p180 kits (Biocrates Life Sciences, Innsbruck, Austria). Samples were prepared and data collected by UPLC tandem mass spectrometry on an API 6500 (Sciex, Framingham, MA) as per manufacturer’s instructions.

### Fat burner classification

The propensity of an individual to metabolically combust (i.e. burn) ingested fat was quantified from the MMCT indirect calorimetry data as both continuous and categorical variables at baseline, 2 and 8 weeks. The continuous variable, % fat burned (%FB), was determined with the following equation:

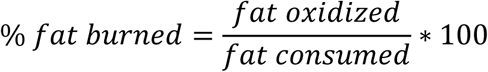

where,

fat oxidized = Frayn equation calculated sum of the 6h PP g fat oxidized
fat consumed = MMCT g fat ingested

Participants were classified based on the magnitude and stability of the % FB: high fat burners (HB) > 40% at all weeks (n = 6); low-fat burners (LB) < 30% at all weeks (n = 7); variable burners (VB) = a mix of HB and LB classification at different weeks (n = 31).

### Calculation of appearance, disappearance, and area under the curve

To quantify the changes in metabolites from fasting to postprandial, a one-compartment oral bolus pharmacokinetic model non-linear curve fit was applied to all lipidomic and metabolomic data to estimate an “appearance” rate (i.e. rate at which the metabolite appeared in plasma), a “disappearance” rate (i.e. rate at which the metabolite disappeared from plasma) and an area under the curve (See S**upplemental Figure 2**). Physiologically, these could indicate changes happening in early postprandial (appearance) and late postprandial (disappearance) states. These data were used in subsequent analyses as described below.

### Statistical tools

An overview of the analysis performed is presented in **Supplemental Figure 3**. All analyses were performed in JMP Pro 17.2 (SAS institute, Cary, NC) or R Statistical software^38^, unless otherwise specified. Data were evaluated for distribution (normality) using Q-Q plots and Shapiro Wilk tests, followed by transformations, if necessary, using the Johnson family of normalizations. Data were evaluated for missingness using the Amelia package in R^39^, and 3% of data were found to be missing. Missing data were imputed using singular value decomposition (SVD) imputation in JMP Pro 16.1, after careful evaluation with several imputation tools (see **Supplemental Figure 4**).

### Univariate statistics

Diet effects on parameter means were tested by analysis of covariance (ANCOVA) with the baseline values (week 0) used as a covariate. Energetic parameters tested included resting metabolic rate (RMR), respiratory exchange ratio (RER), substrate oxidation (carbohydrate and fat oxidation), and metabolic flexibility. Here, we calculated metabolic flexibility as the change in RER and fat oxidation between postprandial and fasting measures (360-0, 180-0, 30-0 mins). To evaluate the effect of the intervention on energy substrate parameters, a repeated measures mixed model was used with diet group, week and time as fixed effects, treatment group and week as an interaction and participant as a random effect, with week as the repeated measure. A two-one sided equivalence test (TOSTER package in R) was used to ensure that metabolic flexibility from the two intervention groups could be combined into a single population for analysis. This analysis tests whether an intervention had a statistically measurable impact on the primary outcome (RER) which is also clinically or physiologically relevant.

FB group differences in postprandial measures of metabolic flexibility, body composition, and circulating lipids over the course of the study were assessed using mixed models. These models included FB-group, week, time (mins) and interaction (FB-group*week) as fixed effects, and participant as a random effect with week as the repeated measure. Area under the curve was calculated based on Simpson’s rule^41^ and Kruskal-Wallis non-parametric tests were used to compare the AUCs between FB-groups by week.

### Multivariate statistics

To enhance interpretation of this highly dimensional data, metabolomic/lipidomic data (∼230 parameters) were subjected to variable clustering using an implementation of the VARCLUS algorithm in JMP Pro v 17.2.0. To highlight the metabolomic features of interest associated with the fat burner group, partial least squares (PLS) analyses were performed using the cluster component scores and either as (a) continuous %FB or (b) as %FB categorical extremes (i.e. HB vs LB) as outcome variables. The non-linear iterative partial least squares (NIPALS) algorithm was used with leave-one-out cross validation to select the number of factors that minimize the Root Mean PRESS statistic. The Q2 (goodness of prediction statistic) and R^2^ (coefficient of multiple determination) for independent and dependent variables were used to evaluate the model fit. Cluster components with a variable importance in projection (VIP) score of > 1 were identified and interpreted as significant explanatory features for the %FB. In addition to running a PLSDA, since we did the MMCT three times (week 0, 2 and 8), the mixOmics package in R was used to do a repeated measures (multi-level) sparse PLSDA (sPLSDA) to extract the loadings, scores, and VIP variables, to compare with the model developed by the NIPALS algorithm. Only VIP variables identified by both approaches were used for final interpretation.

## Results

Forty-four women who were overweight or obese with insulin resistance were included in this study. Participant profiles have previously been published ^27^. Briefly, at baseline the mean participant age was 47.1 ± 9.5, range 21-64 y and mean BMI 32.4 ± 3.9, range 25.2-39.8 kg/m^2 27^. Intervention groups were well matched by age and anthropomorphic data, and diet-dependent changes in body characteristics and metabolic rate measures were not observed (**Supplemental Table 1**)^27^. Linear mixed models identified no significant differences between diet groups in fat oxidation (p=0.47), carbohydrate oxidation (p = 0.53), RMR (p = 0.77), RER (p = 0.50), % change in fat oxidation from fasting (p = 0.46) and % change in RER from fasting (p = 0.72) from minute 0 (before the MMCT) to minute 60, 180 or 360 (see **Figure 1**).

Since diet intervention groups were not statistically different in energy and substrate metabolic parameters and metabolic flexibility, this was further evaluated using equivalence tests to justify combining the intervention groups. The difference between RER from minute 0 (before the MMCT) to minute 360 (6 hours after the test) was used as a primary outcome to evaluate the equivalence of groups. A range of deltas for RER between +/− 0.1 to +/− 0.01 were tested as a change of +/−0.01 constituting a < ∼3% change in RER which was deemed clinically irrelevant based on the American Heart Association report suggesting a 3% within participant measurement variability ^40^. In our study, the maximum measured change in RER between the fasting and postprandial states was 0.294. Results for this test (**Supplemental Figure 5**) concluded that the changes in RER were significantly similar up to our pre-determined delta, and we could proceed in combining the diet groups and assessing all the data together. While this supplemental figure only shows these relationships for week 8, week 0 and week 2 were also tested with identical results.

**Figure 1.**
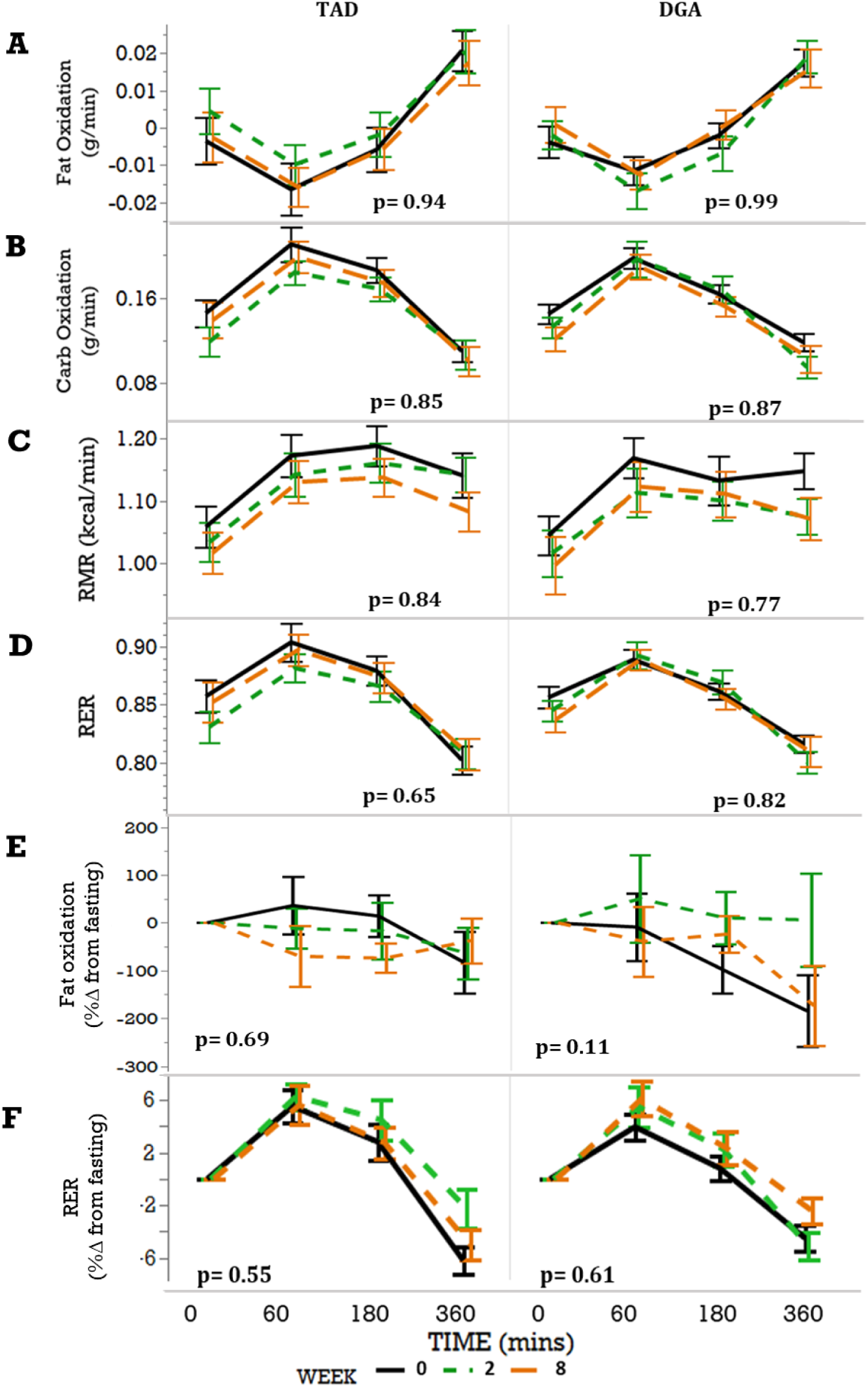
No substantial effect of diet on parameters of metabolic flexibility over the course of the study. Mixed models (with treatment group (TAD or DGA), week, and time as fixed effects, participant as random effect, week as a repeated measure and treatment group by week interaction) were used to evaluate the effect of intervention, and weeks on intervention in (A) fat oxidation rate, (B) carbohydrate oxidation rate, (C) Resting metabolic rate (RMR), (D) respiratory exchange ratio (RER), (E) change in fat oxidation, represented as percentage and (F) change in RER also represented as a percentage in TAD (n = 22) and DGA (n = 22). The p-values inset in the figures indicates the lack of a week effect within that group, and the week x group interaction was not significant in these parameters (data not shown).

Participants were classified into fat-burning groups based on the % of consumed fat from the MMCT burned over the six hours post-test. High burners (HB) burned 40% or more of the ingested fat, while low burners (LB) burned less than 30%. As shown in **Figure 2 panel A**, the VB participants showed inconsistent segregation into a low or high burner group. These characteristics needed to be consistent over the three test weeks of the study to be classified into either group. It is important to note that fat oxidation, RMR and RER were significantly different between burner groups, with HB consistently burning more fat than LB. However, %change from fasting in fat oxidation and RER showed no significant differences between the groups using linear mixed models (Figure 2 panel B). Upon visual inspection, however, it appears that the HB group reduces fat burned as a change from baseline compared to LB at weeks 0 and 8. **Supplemental Figure 6** shows % change from fasting in fat oxidation and RER only in HB and LB groups, where linear mixed model identified a significantly lower % change from fasting in fat oxidation at 60min postprandial in HB compared to LB (p = 0.019).

**Figure 2.**
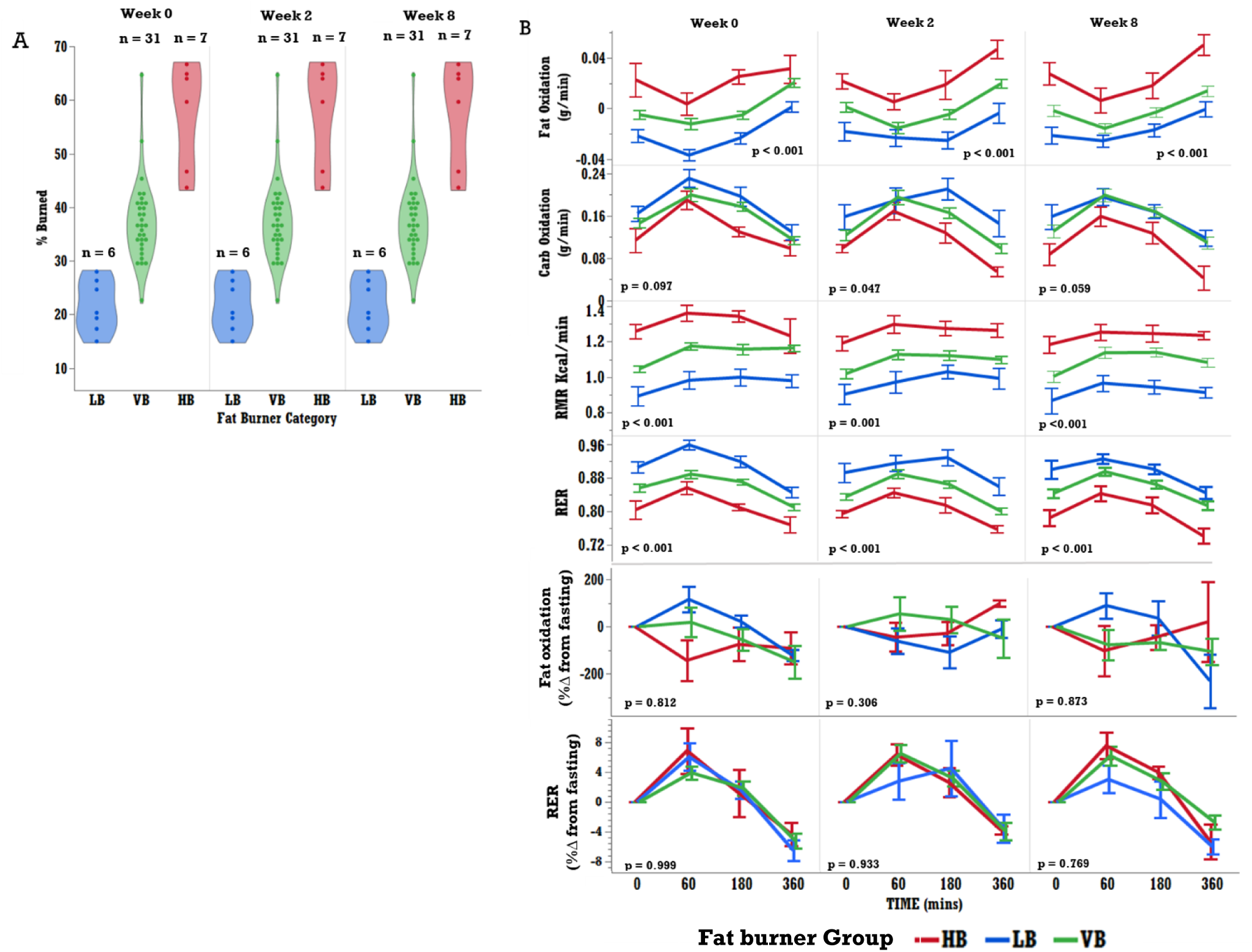
Fat burning groups show significant differences in metabolic features: (A) Fat burning groups, high burners (HB), low burners (LB), and variable burners (VB) were determined based on the % of fat from the challenge meal that was burned over the 6 hours under observation. Participants were separated into fat burner groups (HB burned > 40% of consumed fat, LB burned < 30% of consumed fat) if they sustained their burner classification at weeks 0, 2 and 8 of the intervention. Once categorized, variables relating to metabolic function were assessed by group**. (**B) p-values in figure were derived via mixed model, with fat burning group (HB or LB), fat burning group*time interaction, and time as fixed effects, and participant as a random effect, and time as a repeated measure. The p-values represent group differences in postprandial metabolic response of all FB-groups. HB (n = 6), LB (n = 7), and VB (n = 31).

A summary of clinical outcome variables by fat burner group at baseline and the end of the intervention is presented in **Figure 3**. When separated by %FB group, there were no statistically significant differences in insulin (p = 0.06), glucose (p = 0.61), TG (p = 0.55), HDL (p = 0.11), LDL (p = 0.52), and total cholesterol (p = 0.80) at baseline between HB and LB and this largely persisted through week 8. However, HDLc AUC was significantly lower in HB compared to LB at weeks 2 and 8 (p = 0.03 and 0.01 respectively). Further, while the time course for triglycerides appears visually different between LB and HB in the late postprandial phase, we were not powered to detect the difference statistically. There were significant differences between HB and LB in BMI (**Table 1**) at week 0 (p = 0.023) which persisted through the end of the intervention (week 8 p = 0.023). The same was true for total mass (week 0: p = 0.023, week 8: 0.023), lean mass (p = 0.023, 0.023), fat mass (p = 0.023, 0.023), but not waist to hip ratio (p = 0.49, 0.48) or McAuley’s ISI (p = 0.749, 0.886). There were also no significant group differences for % android fat (p = 0.098, 0.098), % gynoid fat (p = 0.48, 0.86), or age (p = 0.098, 0.098). Menopausal status was also not significantly different between the two groups, evaluated by Fishers exact tests (p = 0.29).

**Figure 3.**
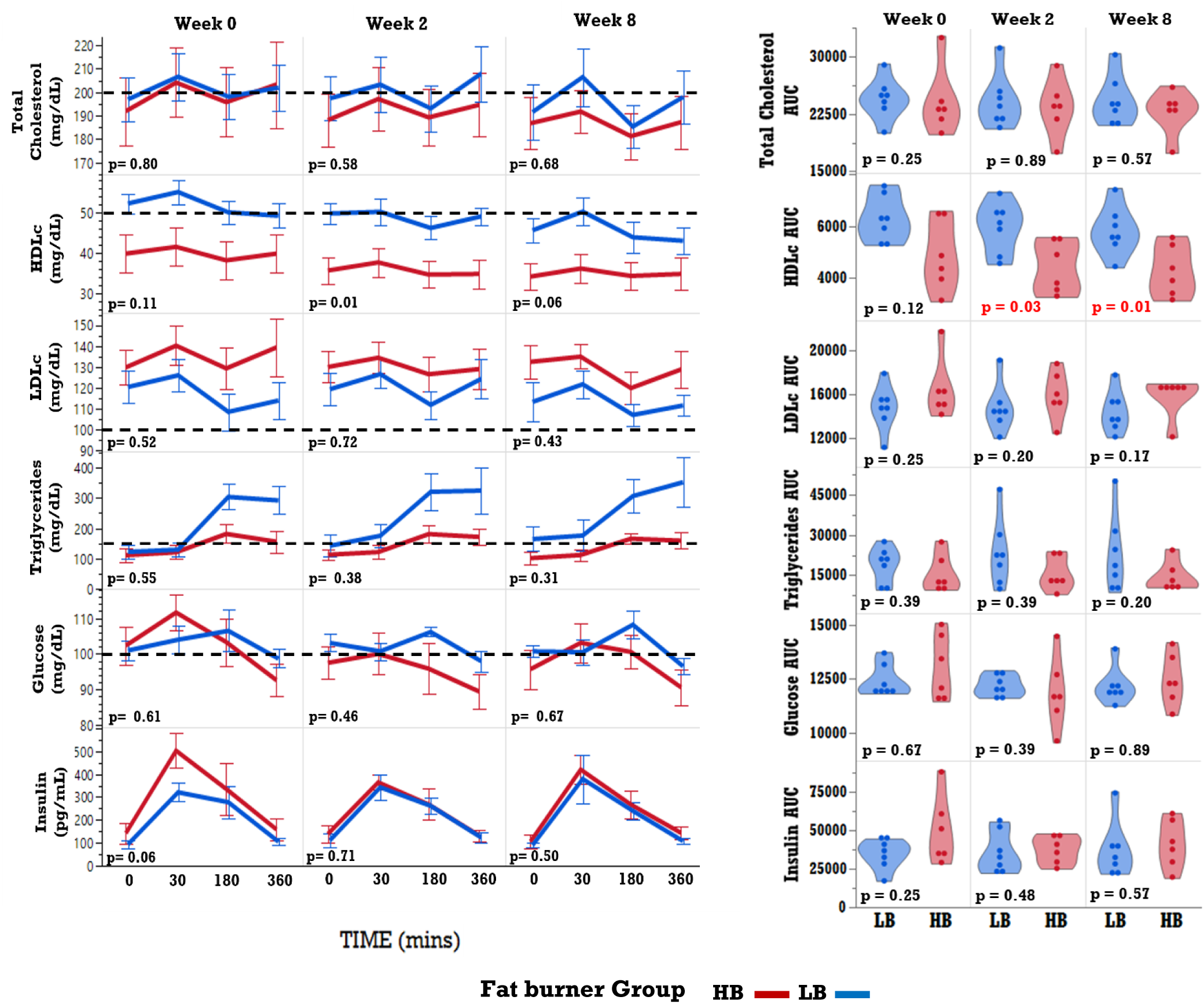
(Left panel) Clinical measures demonstrated subtle differences in key health indicators between fat burner groups: p-values for clinical time course differences over the intervention period were determined through a mixed model approach with fat burner group, and time as fixed effects, participant as random effect, time as repeated measure, along with FB group and time interaction. Horizontal dashed lines represent what is ‘within normal limits’ (WNL for females based on NCEP-STEPIII guidelines) for each measure. Total cholesterol < 200 mg/dL. HDL-c > 50 mg/dL. LDL-c < 100 mg/dL. Glucose < 100 mg/dL (fasted). Insulin < 1010 pg/mL (fasted). p-values reported are between all groups (HB, LB, and VB). (Right panel) Area under the curve, calculated using Simpsons rule for numerical integration, along with p-values inset, based on non-parametric van der Weardan’s tests comparing LB vs HB groups.

**Table 1.** Select anthropometric and clinical measurements at week 0 and week 8 for low fat burner and high fat burner groups.

| Variable | Week 0 |  |  | Week 8 |  |  |
| --- | --- | --- | --- | --- | --- | --- |
|  | LB | HB | p-value | LB | HB | p-value |
|  | mean (SD) | mean (SD) |  | mean (SD) | mean (SD) |  |
| Age | 57.0 (6.4) | 42.7 (13.9) | 0.980 | 57.0 (6.4) | 42. 7 (13.9) | 0.980 |
| BMI (kg/m2) | 29.26 (1.65) | 35.15 (2.96) | 0.023 | 28.67 (2.03) | 34.22 (2.95) | 0.023 |
| Total Mass (kg) | 74.24 (11.79) | 98.67 (9.24) | 0.023 | 72.99 (12.56) | 96.38 (8.08) | 0.023 |
| Fat Mass (kg) | 31.17 (4.80) | 42.53 (4.70) | 0.023 | 41.89 (5.25) | 30.53 (4.84) | 0.023 |
| Lean Mass (kg) | 41.19 (7.07) | 53.81 (4.72) | 0.023 | 40.59 (7.83) | 52.13 (3.45) | 0.023 |
| *Lean mass % | 57.94 (2.50) | 56.95(1.36) | 0.821 | 58.10 (3.46) | 56.62 (1.78) | 0.792 |
| Body Fat % | 42.06 (2.50) | 48.05 (1.36) | 0.280 | 41.90 (3.48) | 43.37 (1.79) | 0.480 |
| Android Fat % of Body Fat | 44.11 (2.37) | 47.87 (3.22) | 0.098 | 43.39 (3.63) | 47.60 (2.55) | 0.098 |
| Gynoid Fat % of Body Fat | 43.31 (3.77) | 44.68 (3.45) | 0.480 | 43.69 (3.72) | 44.70 (4.05) | 0.860 |
| Waist to Hip Ratio | 0.82 (0.03) | 0.86 (0.07) | 0.490 | 0.81 (0.04) | 0.85 (0.06) | 0.480 |
| McAuleys ISI | 9.76 (0.86) | 9.93 (1.24) | 0.749 | 9.71 (1.06) | 9.78 (0.57) | 0.886 |
| Menopausal Status |  |  |  |  |  |  |
| Pre- (count) | 4 | 2 | 0.29** |  |  |  |
| Post- (count) | 2 | 5 |  |  |  |  |

\*lean mass includes bone mineral content
\*\*Fisher's exact test demonstrated no significant association between HB and LB and menopausal status at baseline.

### Variable Clustering dimension reduction

The metabolomic and lipidomic data included 236 variables (not including anthropometric, clinical, and metabolic variables). To better equip our analysis tools to detect metabolite predictors capable of differentiating between the FB groups, we used a dimension reduction algorithm that clusters variables. This dimension reduction tool generates components (like principal components analysis) that are a linear combination of variables. Based on these components, variables are placed into clusters of ‘similar’ variables (cluster components), such that the first cluster component (eigen vectors) within each cluster captures the most variance amongst those variables. In subsequent analyses, the cluster component scores for appearance, disappearance and area under the curve were used as independent predictors to identify differences between the fat burner group metabolic signatures. For appearance rates, AUC, and disappearance rates, clusters of 32, 31 and 36 variables were identified respectively.

### Lipidome predictors of fat oxidation and metabolic flexibility

In our efforts to identify metabolites that were most predictive of %FB groups, only the disappearance rate cluster components (i.e. the 36 variable cluster components that were generated using the variable cluster algorithm in the previous step) resulted in a converged model. **Table 2** lists select clusters and their component metabolites, their corresponding cluster identifier numbers, and eigen vectors (cluster components). Since the eigenvector directions (positive and negative) only translate to the scaled transformations to achieve the clustering, both positive and negative eigen vectors will be interpreted as positive integers in the next steps. The PLSDA models did not converge when the VB were included as an intermediate ordinal group, nor did they converge when we used PLSR to predict % FB as a continuous variable. Thus, our final PLSDA model only compared HB vs LB groups and converged with 2 minimizing factors which explained 17.7% of variation in X and 71.6% of variation in Y variables with a Q^2^ of 0.20 (See **Figure 4**). While the Q^2^ of 0.20 does not suggest complete discrimination between groups, our sPLSDA efforts showed strikingly similar outcomes, and scores and loadings values, indicating the robustness of identified differences. The scores plot of the PLSDA, inset into the loadings plot, shows only a small overlap between HB (in red) and LB (in blue) groups. The loadings plot, which depicts the corresponding cluster components shows which variables brought about the separation. Clusters with VIP > 1 are semi-synonymous to those with p-values < 0.05, and therefore play a significant role in differentiating the HB and LB group. In addition, the results from the sPLSDA are presented in **Supplemental Figure 7** and show very similar separation of participants by burner classification, and VIP variables, with a total of 17% of X was explained by the first two loadings, like the PLSDA using NIPALS algorithm.

**Figure 4.**
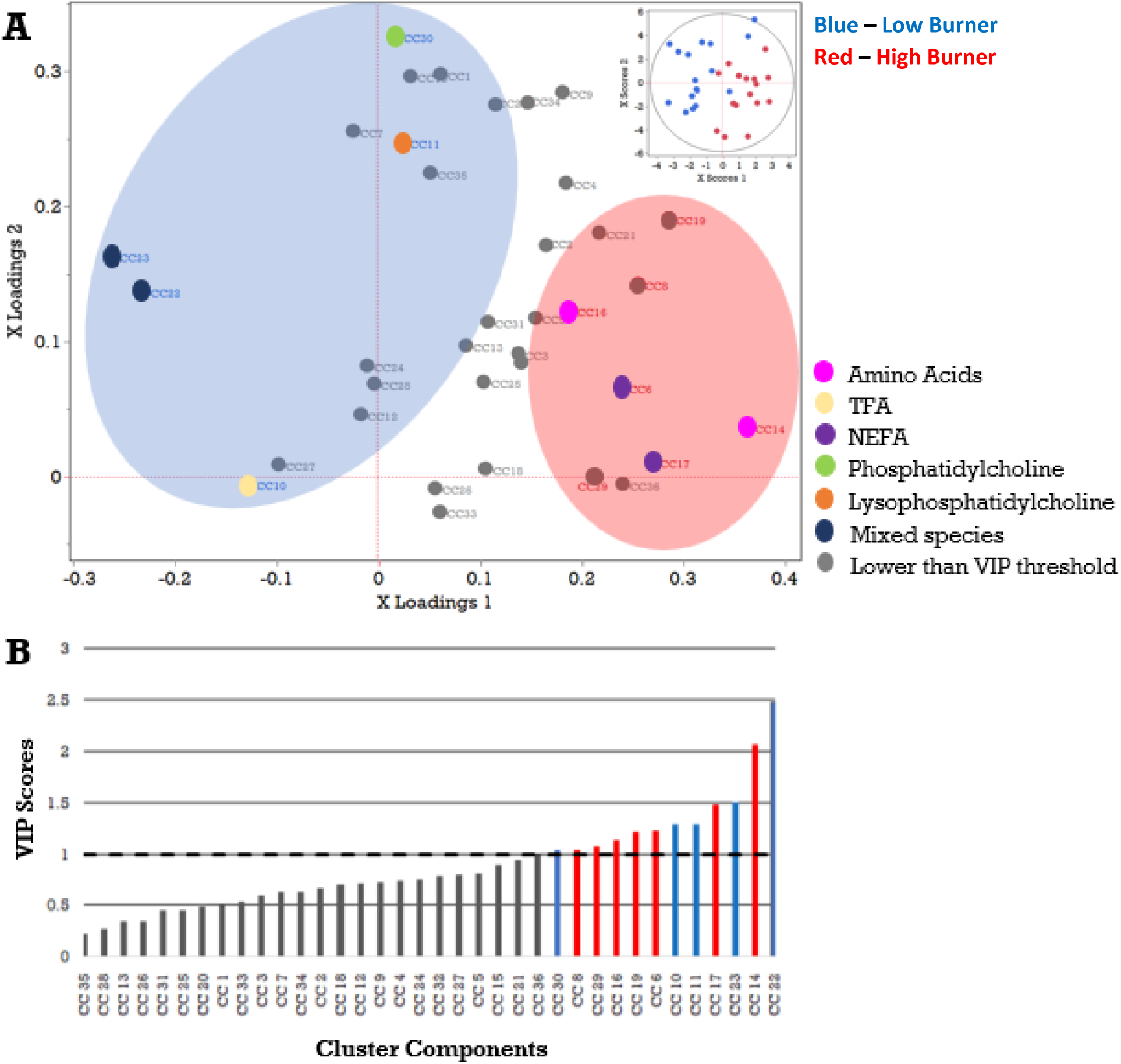
PLSDA demonstrates clustering of various lipids that are strongly associated with fat burning groups. Cluster components (CC) from variable clustering of disappearance rates of metabolites were used as input predicting fat burner groups HB and LB. Inclusion of VB did not allow for model convergence, so the presented model is only comparing HB and LB. **(A)** Highlights the clusters that were associated with each burner group. HB (n = 6), LB (n = 7), and VB (n = 31). Partial Least Squares Discriminant Analysis using NIPALS algorithm converged on a model with 2 minimizing factors, where 17.7% of X and 71.6% of Y variables were explained with a Q^2^ of 0.20 **(B)** Variable importance plot (VIP) scores were chosen for analysis if they were over 1, with blue indicating association with LB and red for HB. TFA: Total Fatty acids, NEFA: Non-esterified fatty acids

**Table 2.**
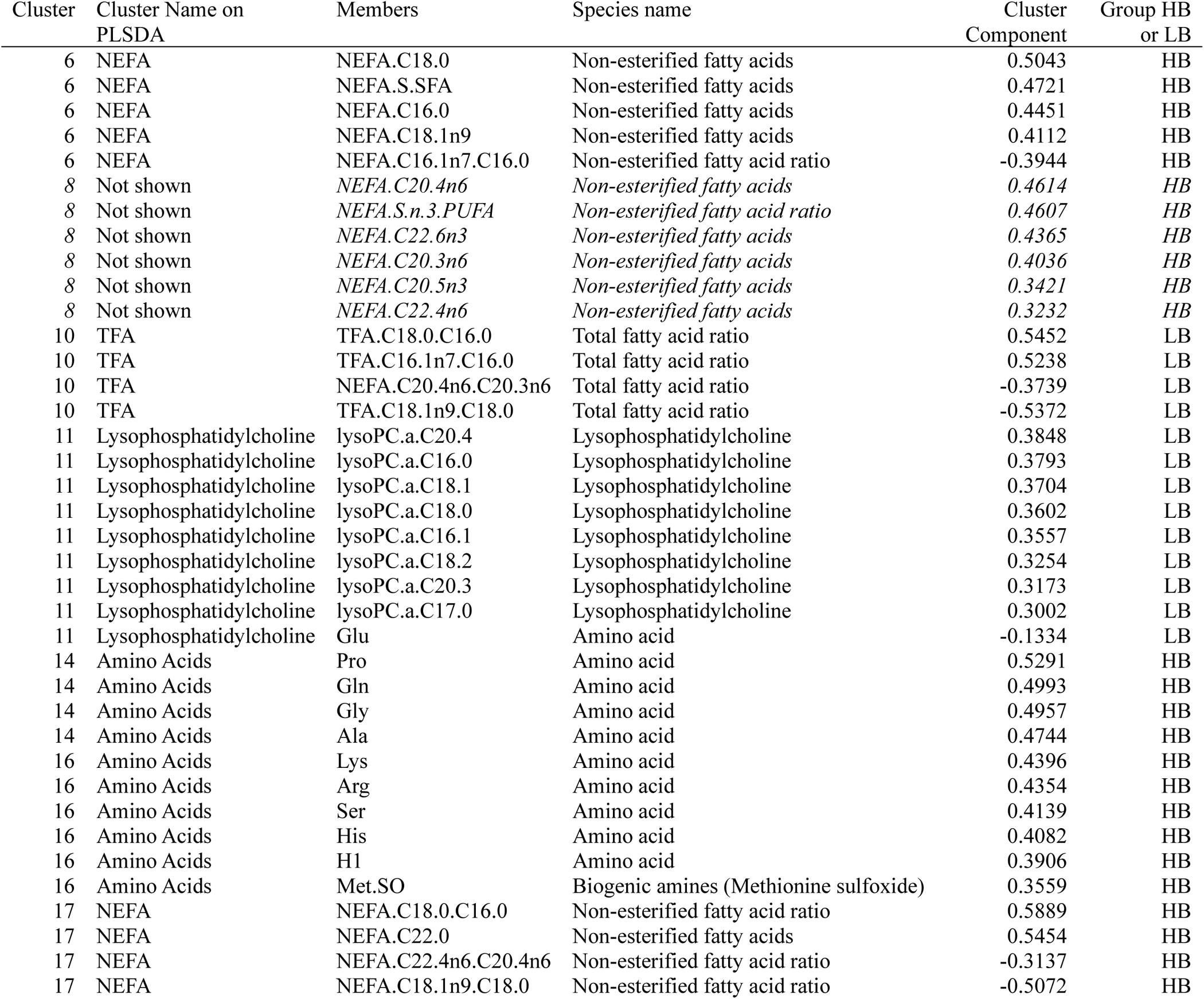
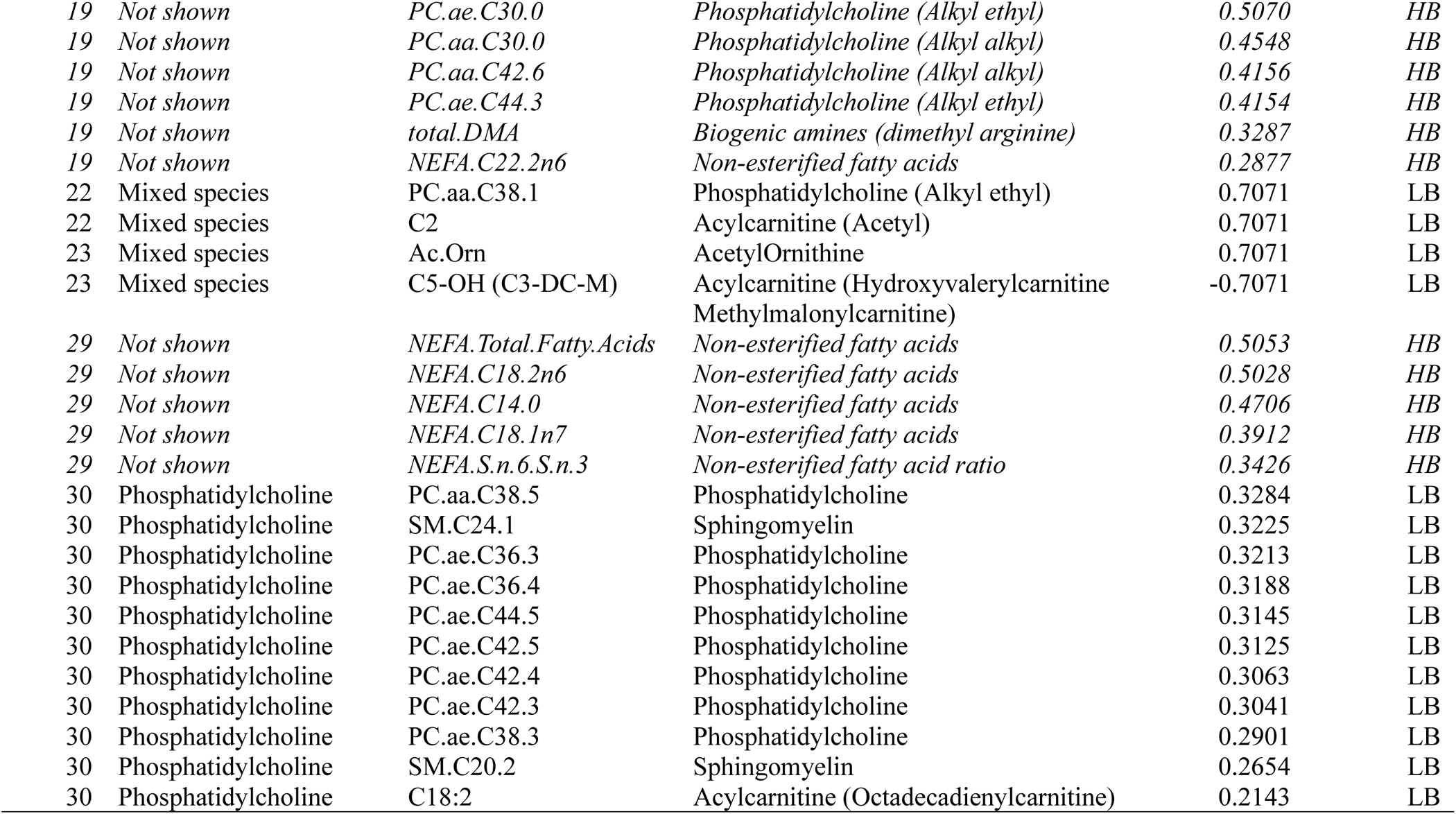
List of each metabolite by cluster for the Partial Least Squares Discriminant Analysis (PLSDA).

**Table 2** lists all the VIP variables from the PLSDA and sPLSDA analyses. Only the commonly identified cluster components from both analyses will be used for interpretation. As shown in Figure 4 panel B, cluster components 10, 11, 22, 23 and 30 were predictors of the LB group, while components 6, 14, 16 and 17 were predictors of the HB group.

#### LB group metabolites

As seen in Table 2, cluster component 10 includes higher (faster) disappearance rates from plasma of total and non-esterified fatty acid ratios TFA C18:0/16:0, TFAC16.1n7/C16.0 (C16/18 elongase and Δ9-desaturase), NEFA C20.4n6/C20.3n6 and TFAC18.1n9/C18.0. Cluster 11 includes lysophosphatidycholines (LPC) with long chain saturated, monounsaturated, and polyunsaturated fatty acids. Cluster 22 and 23 are a mixture of acylcarnitines (acetylcarnitine and methylmalonylcarnitine), acetylornithine, and phospholipids (PC.aa.C38.1). Cluster 30 is a mixture of acylcarnitines (octadecadienylcarnitine), sphingomyelins and alkyl-ethyl phosphatidylcholines.

#### HB group metabolites

Cluster 6 includes higher (faster) disappearance rates from plasma of total NEFA saturated and monounsaturated fatty acids and the ratio of C16:1n9/C16. Cluster 14 and 16 include all non-essential amino acids, while cluster 17 includes non-esterified fatty acid ratios (C18:0/C16:0, C22:4n6/C20:4n6 and C18:1n9/C18:0).

## Discussion

In the current study, as expected, there was no differential effect of the diet interventions on our chosen measures of metabolic flexibility. Further, we identified individuals who were consistently different in how they handled the influx of energy substrates from the MMCT i.e. high fat burners (HB) and low-fat burners (LB). Women in the HB group consistently oxidized >40% of consumed fat, while the LB group consistently oxidized < 30% of consumed fat. Based on what is understood about fat oxidation, metabolic flexibility, and insulin resistance, overall low-fat burning is associated with reduced metabolic flexibility^2^. However, in our cohort, women who burned less fat at fasting (LB group) switched to burning more fat during the 6h postprandial, and in the immediate postprandial phase after consuming the MMCT (with 60% fat, 28% carbohydrates and 12% protein) compared to the HB group. Our sample size was likely too small to detect this at week 2 and week 8, but did identify this difference at week 0, and can be visually seen at week 8. Further, the LB group had lower fat mass, lean mass, BMI and higher HDL compared to the HB group. However, proportions of lean and fat mass were not significantly different between groups. This suggests that in cases of high BMI and insulin resistance, matched proportional lean mass does not “rescue” the effect higher fat mass has on metabolic flexibility^42^. While women in the LB group were older and more postmenopausal, the HB group was not homogenously younger or premenopausal. While metabolic flexibility differences at week 0 were observed in the early postprandial phase, metabolomic and lipidomic profiles identified higher late-postprandial disappearance rates of lysophosphatidylcholines and acylcarnitines in LB group, both implying better metabolic health.

Women in the DGA group were given more whole fruits, vegetables, whole grains, seafood, and nuts, whereas women in the TAD group were given more refined grains, meat, and solid fats over the 8-week feeding intervention^27^. As predicted, there was no impact of this diet on metabolic flexibility, measured by RER or change in fat oxidation between fasting and postprandial time points. Similarly, Kardinaal et al.^43^ evaluated RER changes in a group of healthy males given a high-fat meal over 4 weeks and did not see any changes in their RER. Likewise, in Fechner et al, a group of males and females randomized to six weeks of either a healthy diet or western diet, also did not experience changes in insulin sensitivity or RQ when measured by high fat meal challenge test^19^.

In the late postprandial phase, there was an increased rate of disappearance of Lysophosphatidylcholine (LPCs) and phosphatidylcholine (PCs) in the LB group of women when compared to HB. LPCs are phosphatidylcholines that have been cleaved by a phospholipase (see **Figure 5**), and more commonly function as lipid mediators^44^. The composition of both PC and LPC are primarily saturated fatty acids (SFA) or monounsaturated fatty acids (MUFA) with varying degrees of length and position of unsaturated double bonds. While PCs are found in membranes in large quantities, higher concentrations of LPC have been associated with atherosclerosis through disruptions of mitochondrial integrity^44^, while lower circulating concentrations are found in prediabetes and type 2 diabetes^45^ particularly in the skeletal muscle^46^. LPCs are known to inhibit hepatic fatty acid oxidation (among several other effects)^47^. Hence, their clearance would have supported maximal fat oxidation in LB. LB women could have reduced plasma phospholipase2 (PLA2) activity, which generates LPC from PC. Higher PLA2 activity has been observed to be proatherogenic, irrespective of whether this is in plasma or in the endothelium^48^. So, this higher rate of disappearance of LPC could be a largely metabolically favorable observation. Further, this suggests that relatively quicker disappearance of plasma LPC’s following a high fat meal could be indicative of higher metabolic flexibility.

**Figure 5:**
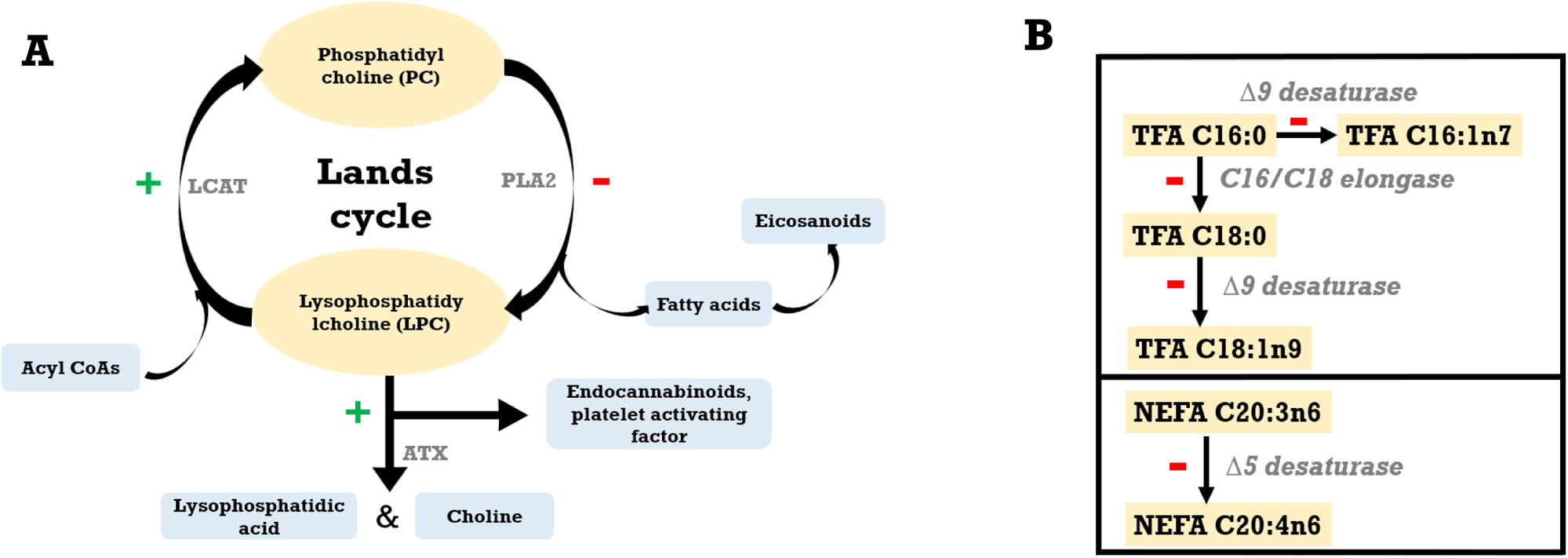
A summary of late postprandial (disappearance) metabolite changes that delineated HB and LB phenotypes. A and B summarize metabolic milieu in LB and C and D do the same in HB group women. The green ‘+’ and red ‘-‘signs denote the speculated increase and decrease of metabolic processes based on observed disappearance trends. Metabolites in yellow oval/boxes were measured parameters, and light blue boxes are upstream or downstream metabolites not measured and hypothesized based on known biosynthesis and degradation pathways.

LB women also displayed a faster rate of disappearance of acylcarnitines from plasma. Higher circulating concentrations of acetylcarnitine have been reported in people with prediabetes and type 2 diabetes^49^. Also, increased circulating octadecadienoylcarnitine has been associated with increased all-cause mortality and hospitalizations in heart failure patients^49^. Together, these would suggest that a quicker disappearance of from plasma in LB women would lead to a metabolically healthier milieu compared to women in HB group.

In the late postprandial phase, especially following a meal that has carbohydrates in it (which our MMCT did), the initial rise followed by the post-absorptive drop in insulin concentration will result in an increase in adipose lipolysis and higher circulating NEFA concentrations^50^. In this phase, the LB group had a reduction in Δ9 desaturase, Δ5 desaturase and C16/18 elongase activities affecting the complex lipid pool (fatty acids attached to triglycerides) likely resulting in more C16:0 and C18:0 acyl glycerol concentrations in plasma. Saturated TGs, such as C16:0 and C18:0, have been shown to suppress LDL-receptor activity, thereby increasing plasma LDL-c^51^, increasing cardiovascular disease risk. Thus far, this is the only less-than-ideal metabolic milieu that has been noted in women in the LB group.

Our initial assumption was that those who could burn a higher percentage of fat following the challenge meal would be more metabolically flexible, as they would demonstrate an effective switch of primary metabolic fuel sources, potentially leading to health benefits. However, our findings in this cohort do not support this idea, suggesting a more complex relationship between fat oxidation and metabolic flexibility. HB women had higher body mass and were in the obese BMI category compared to the LB group who were in the overweight BMI category. Obese insulin resistant individuals are known to have higher systemic fasting respiratory quotient^2,52^ but this is not true in our current cohort. Metabolic inflexibility to a high fat meal has been shown to be a predictor for subsequent weight gain^17^, suggesting our HB group women are metabolically inclined to further gain body weight. Differences in their underlying physiology detected by omic analysis, such as faster clearance from circulation of LPCs and acylcarnitines to offset the higher saturated triglycerides, also support our observation that LB women may be on the “healthier” metabolic spectrum compared to HB women. Given our findings of higher fat burning not equaling higher metabolic flexibility, the definition of metabolic flexibility may need to be expanded beyond the ability to burn fat in a high-fat challenge meal. A follow up study that evaluates how much fat oxidation and fuel switching occurs following consumption of meals with serial increasing % dietary fat (0 – 100%) in a diverse population could shed further light on this relationship.

### Strengths and Limitations

This investigation used a standard meal challenge test for all the women and repeated it three times, which affords robustness to our conclusions. Participants were categorized into burner groups based on consistent responses in all three challenge events, reducing the likelihood of measurement errors driving our sub-group determination. All participants were relatively sedentary, did not change physical activity during the intervention, and were given a pre-test dinner the night prior to consuming the MMCT to reduce variability from pre-meal dietary sources. Evaluating equivalence and robust approaches to data analyses (such as including a repeated measures PLSDA) also strengthen our interpretations. However, there we several limitations that must be acknowledged. Our categorized burner groups were small (HB, n = 6 and LB, n = 7). This small sample size may have been the reason why only one model PLSDA converged among several we tested. It is possible that the difference between the high fat burners and low-fat burners may be due to a genetic predisposition to be obligatory fat or carbohydrate burners, rather than the flexibility or inflexibility of their metabolism. However, the study lacks SNP/genome sequence data that could corroborate this. The high inter-test and inter-individual variability in our VB group (n = 31, ∼70% of participants) unfortunately rendered the group empirically inexplicable. The individuals in the VB group, however, are likely to be the majority in a population, highlighting the importance of being able to understand their underlying physiology. Future studies must use larger sample sizes to be able to overcome this failing. While we controlled the participants’ food intake and physical activity, we did not control or account for changes in sleep, endocrine factors, stress, gut microbiome etc. which could have affected one of the three weeks’ MMCT responses of women in the VB group.

## Conclusion

In summary, we identified that a high fat burning capacity is unlikely to equal higher metabolic flexibility universally in women with insulin resistance. In addition, the HB group seemed to reap no clear metabolic or clinical rewards from this high burning capacity. Currently, there is a lack of consensus on the definition and standard metric for metabolic flexibility, particularly in the way it relates to dietary fat-induced metabolic flexibility. To fully understand the response of the metabolism to gauge flexibility, a mixed macronutrient challenge test offers more information than a standard oral glucose tolerance test. Additionally, we also highlighted the important inclusion of metabolomics, allowing investigators to “peer under the hood” of an individual’s metabolism, corroborating results observed both clinically and physiologically. This may be especially true in insulin resistant individuals, as the relationship between insulin and lipid metabolism is complicated by the inherent involvement of body composition, dietary macro-nutrient composition, and several other endocrine and molecular mechanisms at play. As mentioned by other researchers, a comprehensive definition of metabolic flexibility to lipids is sorely needed, especially given the results mentioned above.

## Data Availability

All data produced in the present study are available upon reasonable request to the authors.

## Author Contributions

Conceptualization SK, NLK, JWN; Methodology SK, NLK, JWN; Formal Analysis SK, VMA, MMA; Investigation NLK, SK, VMA, MMA; Data Curation SK; Writing – Original Draft VMA, SK, MMA; Writing – Review & Editing SK, NLM, JWN, MMA; Visualization SK, MMA; Supervision SK; Funding Acquisition SK, NLK, JWN.

## Declaration of Interests

The authors declare no competing interests.

Declaration of generative AI and AI-assisted technologies: SCITE and Elicit were used to search for references to ensure comprehensive literature review to use in the writing of this manuscript. However, no AI-assisted text was incorporated, and all writing was done by the authors.

## Supplemental Information

Document S1. Figures S1-S7 and Tables S1

## Funding information

National Dairy Council, Campbell Soup Co., USDA-ARS Projects 2032-51530-022-00D and 2023-51530-025-00D. We acknowledge the West Coast Metabolomics Center Pilot and Feasibility study to N.K. and S.K. funded through National Institutes of Health Grant U24 DK097154.

## Clinical Trials Information

Clinicaltrials.gov: NCT02298725

## Notes

### Competing Interest Statement

The authors have declared no competing interest.

### Clinical Trial

NCT02298725

### Funding Statement

This study was funded by National Dairy Council, Campbell Soup Co., USDA- ARS Projects 2032-51530-022-00D and 2023-51530-025-00D. We acknowledge the West Coast Metabolomics Center Pilot and Feasibility study to N.K. and S.K. funded through National Institutes of Health Grant U24 DK097154.

### Author Declarations

IRB of United States Department of Agriculture, Agricultural Research Service, Western Human Nutrition Research Center gave ethical approval for this work.

