## Supplemental Files for "Fat burning capacity in a mixed macronutrient meal protocol does not reflect metabolic flexibility in women who are overweight or obese"

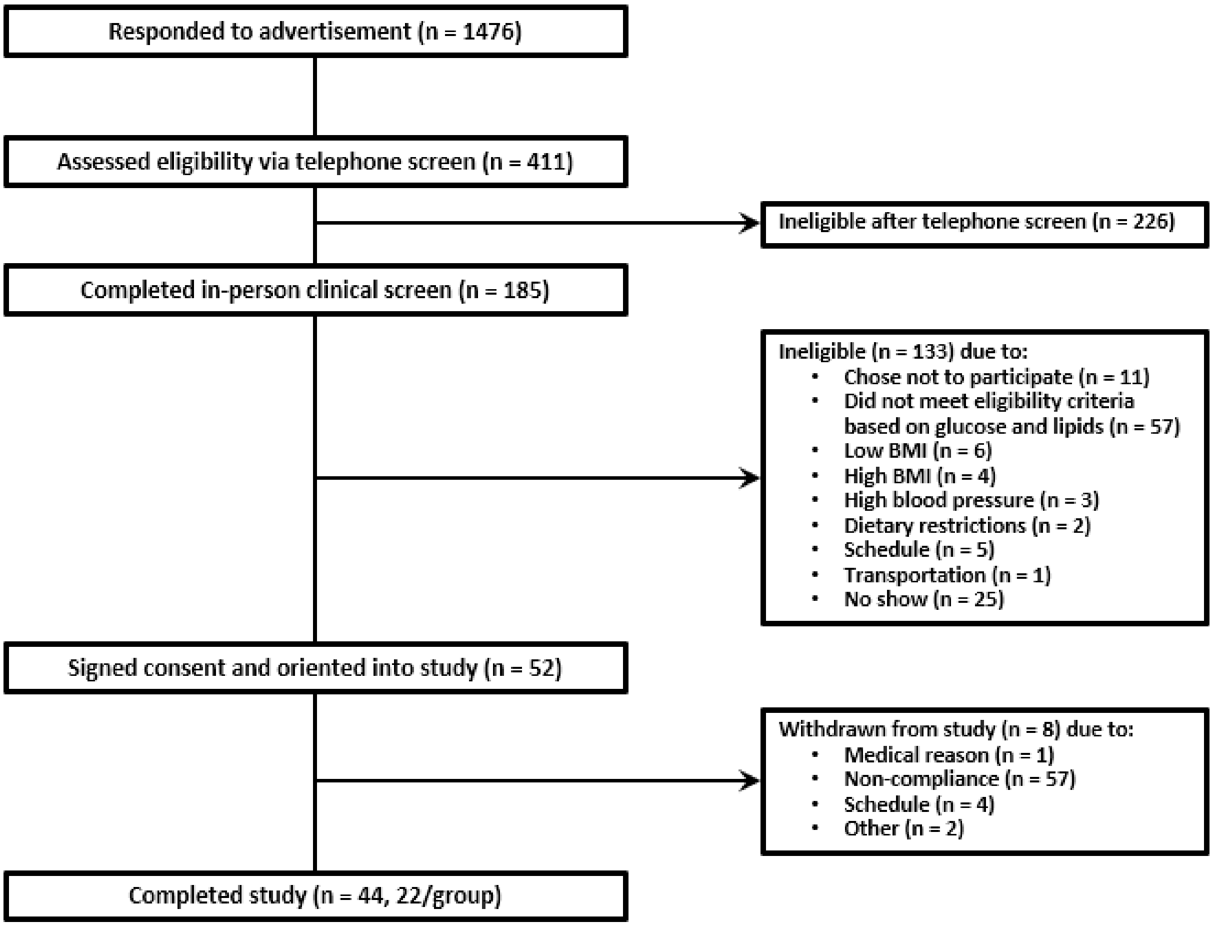


**Supplemental Figure 1. Consort diagram** representing the volunteers who were screened, consented, and completed the study. In the current study completed study volunteers were used in analysis. *Reproduced with permission.*

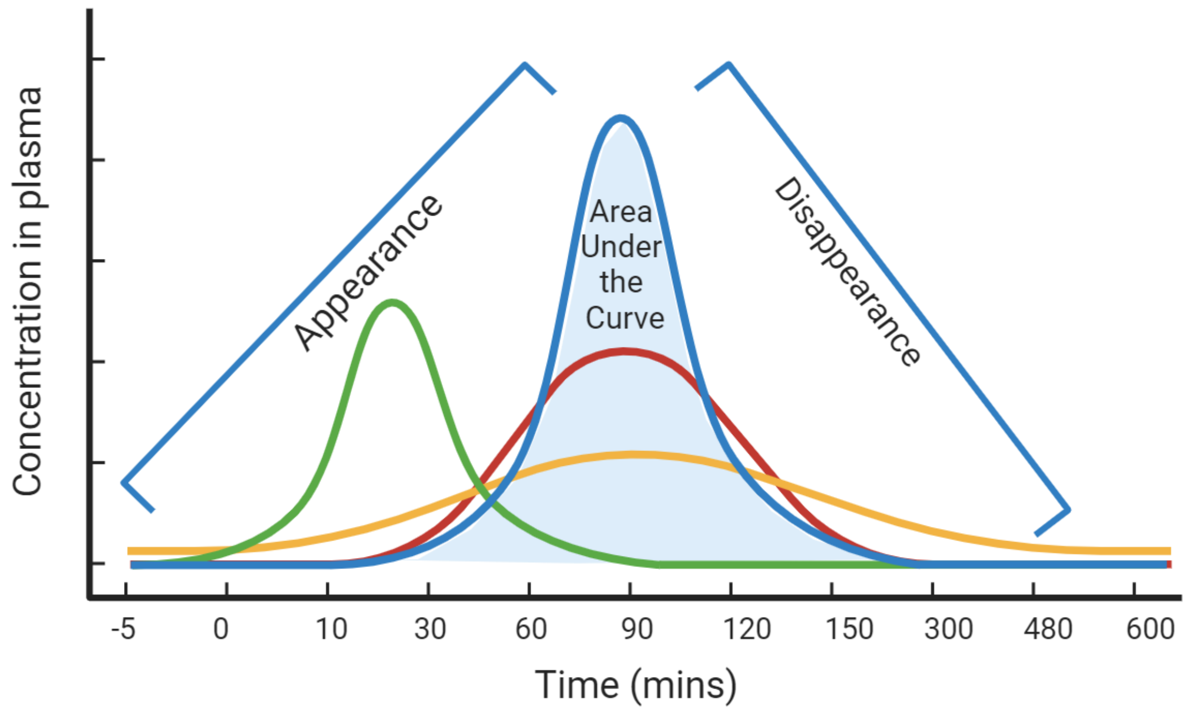


**Supplemental Figure 2.** Figure demonstrating the appearance, area under the curve, and disappearance of metabolites from the blood. Lines in green, red, yellow, and blue depict different patterns of concentration changes of metabolites or lipids in plasma over time. Each curve would have its own appearance, disappearance and AUC computed accordingly.

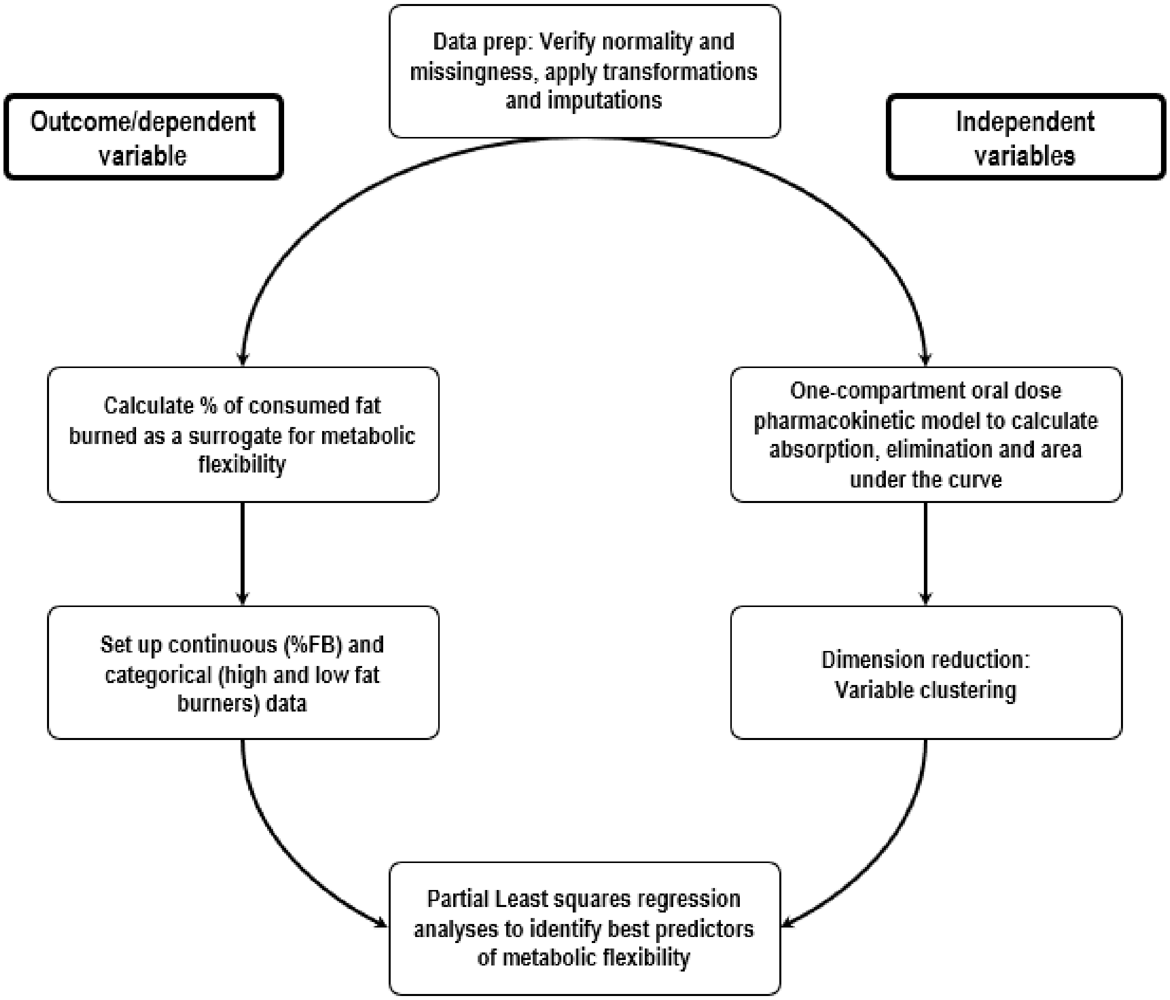


**Supplemental Figure 3:** An overview of the analysis approach used in the presented manuscript. Briefly, the outcome variables were generated as a function of the total g of fat oxidized (measured using indirect calorimetry) and total g of fat consumed (as part of the mixed macronutrient challenge test). The independent variables (metabolomic and lipidomic data) were pre-prepped by transforming to achieve normality, imputed to treat missingness. Since this included fasting and postprandial time course data, a non-linear curve fitting step was used to generate single values to represent the rate at which metabolites appeared (absorption), stayed (area under the curve), and disappeared (elimination) in/from plasma. A dimension reduction algorithm clustered variables to make this very wide dataset usable. These clustered variables were then used in partial least square regression analyses to identify discriminants.

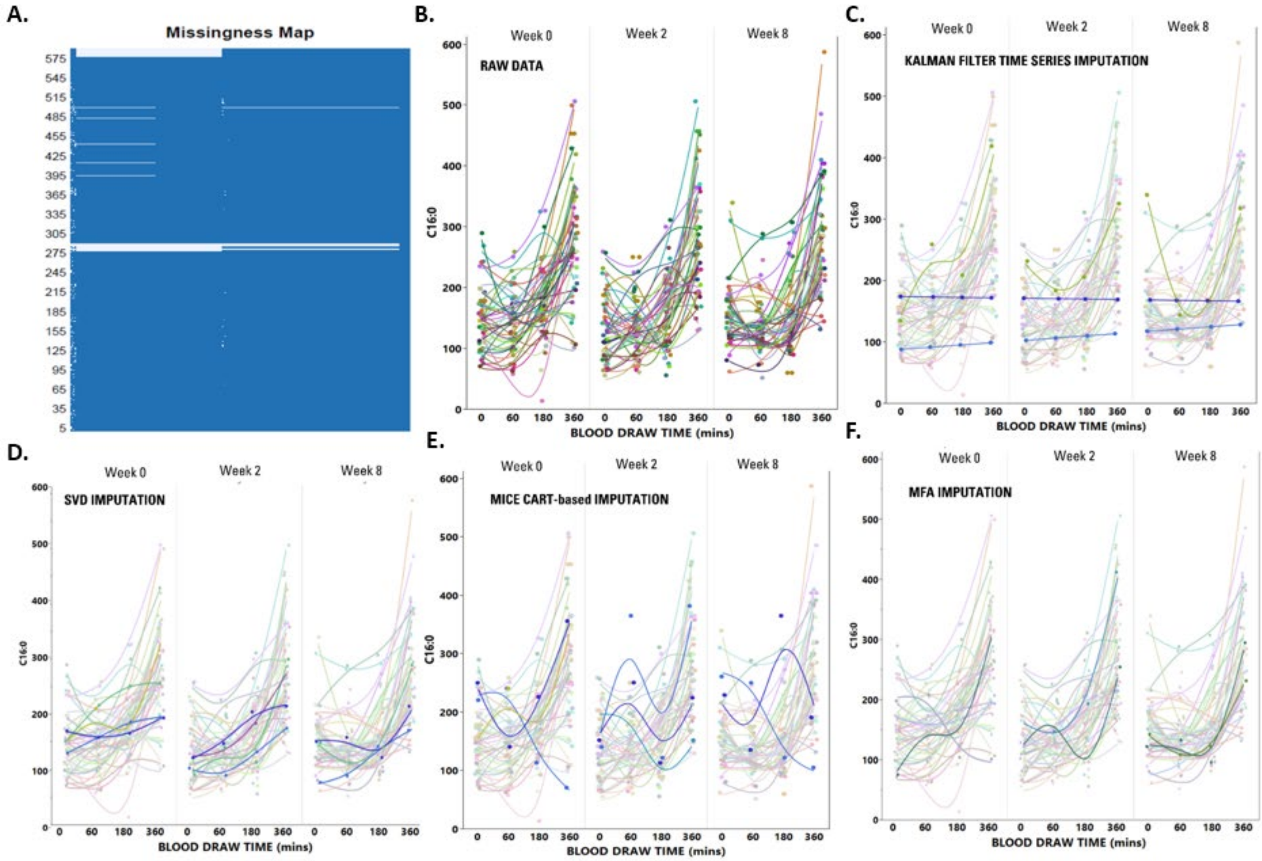


**Supplemental Figure 4**: A total of 3% of total data was missing and deemed suitable for imputation approaches **(A)**. Several imputation techniques were applied to the data including MFA (Multiple Factor Analysis) , MICE CART (Multivariate imputation by chained equations classification and regression trees), Kalman Filter and SVD (singular value decomposition) imputation. Among all chosen tools SVD based imputation **(D)** appeared to perform the best, as demonstrated above for the C16 metabolite. This tool outperformed other options such as Kalman Filter **(B)** which was unable to demonstrate significant time-course changes resulting in little variation in the imputed values. Values imputed with the MICE CART imputation resulted in little changes to the spread of the data with the medians of the raw data and imputed data remaining fairly close at 163.55 and 161.84, respectively.

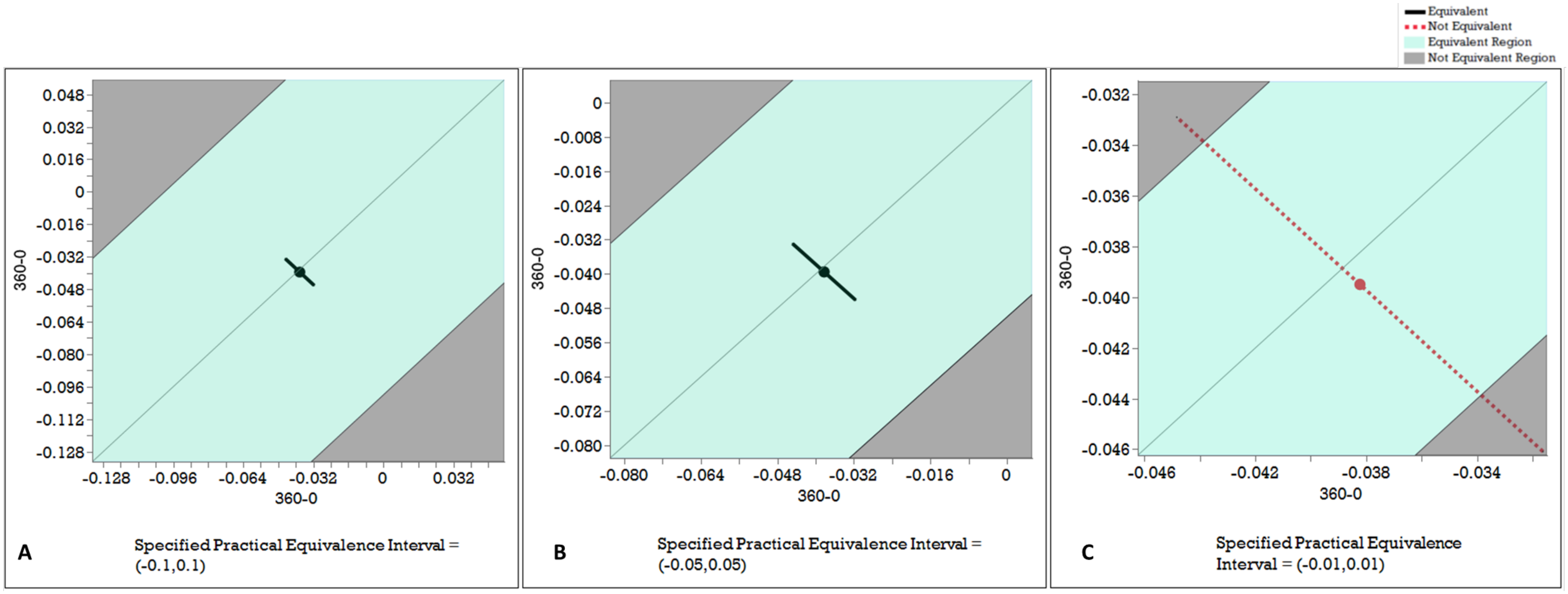


**Supplemental Figure 5**: Non-equivalence test results of RER in the study difference between diet intervention groups shown here at Week 8. Similar analyses were completed for Weeks 0 and 2 with identical results. Panels A and B indicate that a difference of 0.1 or 0.05 in RER were not adequate to be found as non-equivalent, and panel C indicates that a 0.01 interval would be considered non-equivalent. Considering both measurement limitations of a metabolic cart, and physiological changes that would indicate changes in substrate oxidation, a 0.01 change in RER would be impractical to use as a threshold to identify difference between two groups, such as in this case between TAD and DGA.

**Supplemental Table 1**: Metabolic variables evaluated at week 8 by ANCOVA using week 0 as a covariate (by diet). No significant differences were identified between the two groups.

| **Parameter** | **Diet** | |
|---|---|---|
| | **TAD (n = 22) (mean (SD))** | **DGA (n = 22) (mean (SD))** |
| Body weight (kg) | 88.75 (13.23) | 88.60 (15.63) |
| BMI (kg/m2) | 32.83 (3.76) | 31.96 (4.00) |
| Total Physical activity energy expenditure (kcal per week) | 518.8 (177.1) | 505.4 (209.8) |
| Waist circumference (cm) | 95.35 (9.18) | 95.90 (10.33) |
| Hip circumference (cm) | 115.34 (8.73) | 117.12 (10.62) |
| Waist to Hip Ratio | 0.84 (0.06) | 0.82 (0.07) |
| BMC (g) | 2230.18 (370.54) | 2185.62 (442.41) |
| BMD (g/cm2) | 1.15 (0.12) | 1.11 (0.13) |
| Fat Mass (kg) | 37.95 (7.45) | 37.8 (7.70) |
| Lean Mass (kg) | 47.10 (6.71) | 46.95 (7.95) |
| Total Mass (kg) | 87.28 (13.09) | 86.92 (15.33) |
| Trunk % Fat | 44.25 (5.02) | 45.28 (3.67) |
| %Body Fat | 43.30 (3.86) | 43.30 (3.02) |
| Android Fat Mass (kg) | 3.31 (0.95) | 3.48 (1.04) |
| Android % Fat of Body Fat | 45.94 (5.48) | 46.96 (4.55) |
| Gynoid Fat Mass (kg) | 6.43 (1.46) | 6.37 (1.62) |
| Gynoid % Fat of Body Fat | 45.26 (4.61) | 44.71 (4.18) |
| McAuley's Insulin Sensitivity Index (<5.8 cut off for insulin sensitivity) | 10.03 (1.25) | 9.89 (1.16) |
| $VO_2$ L/min | 0.22 (0.03) | 0.22 (0.03) |
| $VCO_2$ L/min | 0.18 (0.02) | 0.19 (0.02) |
| Respiratory exchange ratio (RER) | 0.86 (0.06) | 0.86 (0.05) |
| Resting Metabolic Rate (kcal/day) | 1501.0 (214.1) | 1526.3 (220.4) |

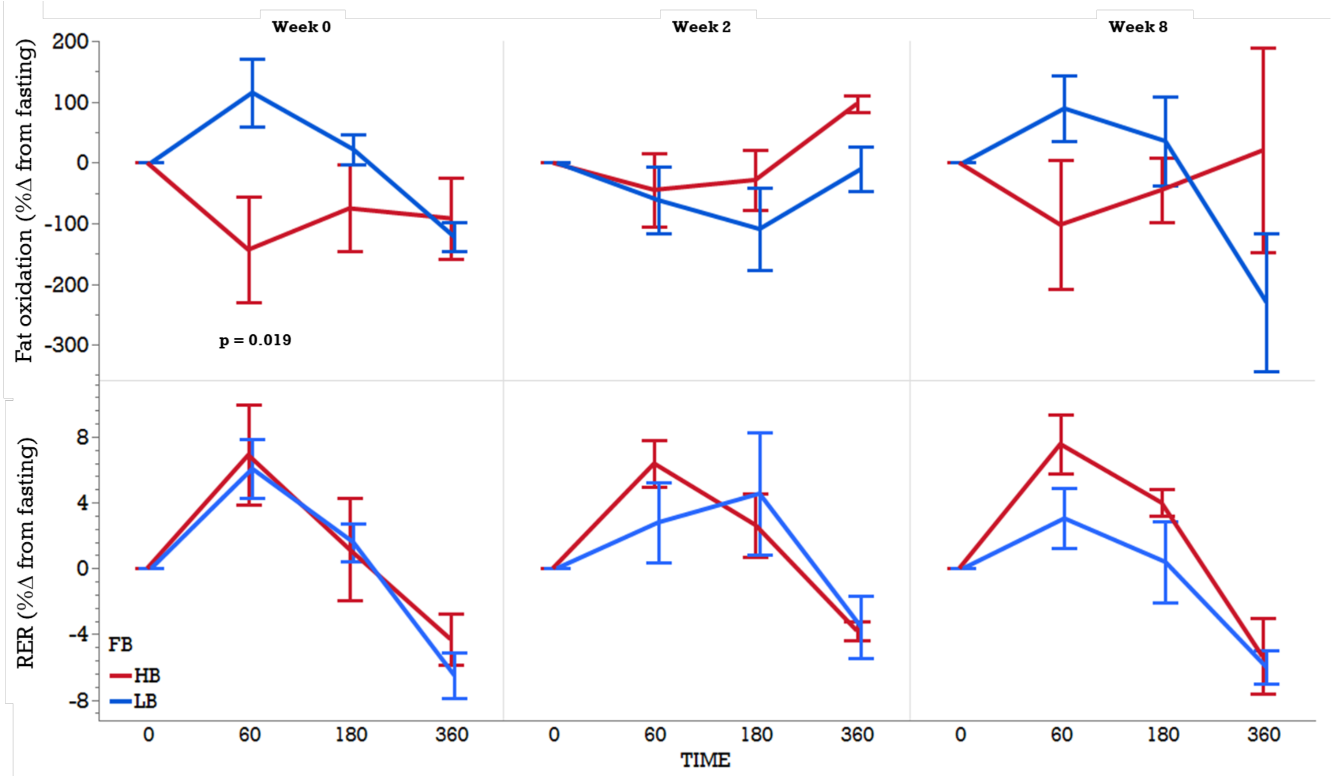


**Supplemental Figure 6:** Comparing only HB and LB groups without VB group in the linear mixed models using burner group, weeks, and time (mins) as fixed effects, week as repeated measures, and participant as random effect identified a burner group by week and burner group by time effect in % change in fat oxidation from fasting, but no differences were identified in % change in RER from fasting. At 60min postprandial, at week 0, HB demonstrated a significantly lower change in fat oxidation in response to the meal test when compared to the LB group ($p = 0.019$). While not detected statistically (likely due to a small sample size), visually we can still see separation between the groups at 60min postprandial in week 8.

**A**

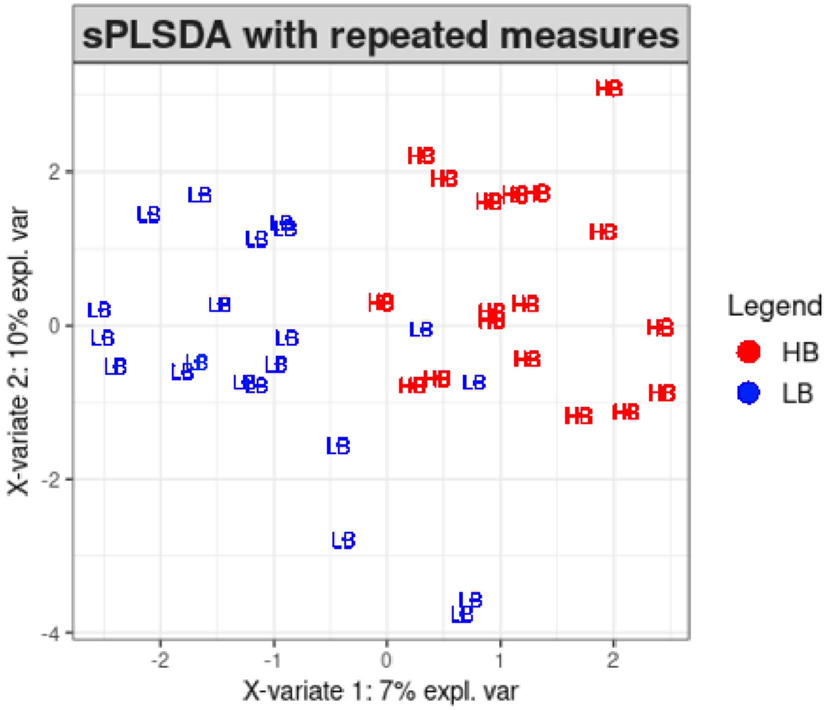
sPLSDA with repeated measures
X-variate 2: 10% expl. var
X-variate 1: 7% expl. var
2
0
-2
-4
-2
-1
0
1
2
HB
LB
Legend
HB
LB

**B**

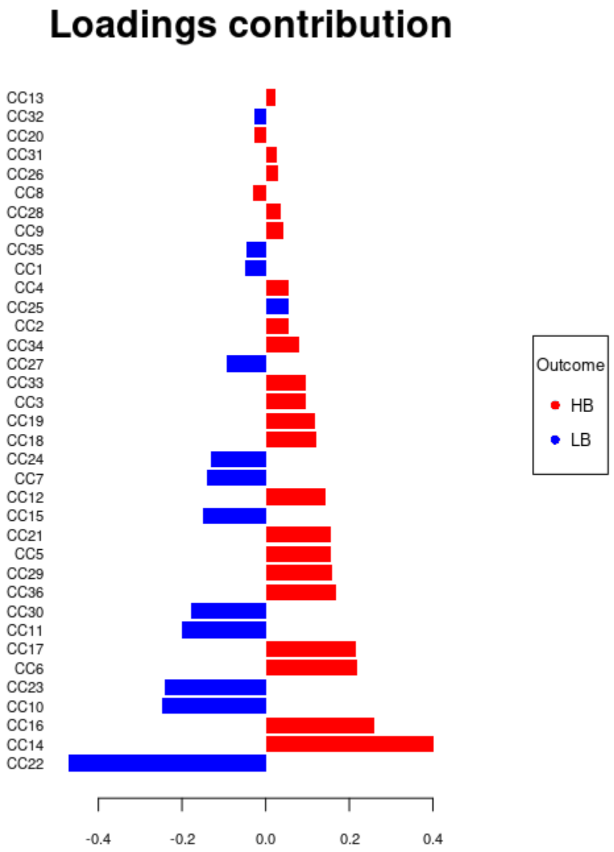
Loadings contribution
CC13
CC32
CC20
CC31
CC26
CC8
CC28
CC9
CC35
CC1
CC4
CC25
CC2
CC34
CC27
CC33
CC3
CC19
CC18
CC24
CC7
CC12
CC15
CC21
CC5
CC29
CC36
CC30
CC11
CC17
CC6
CC23
CC10
CC16
CC14
CC22
-0.4
-0.2
0.0
0.2
0.4
Outcome
HB
LB

**C**

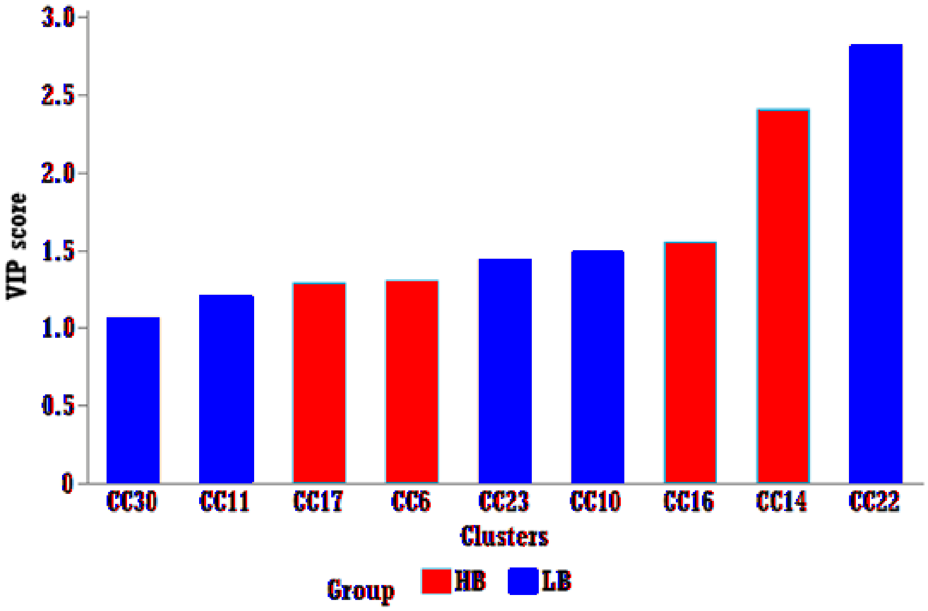


**Supplemental Figure 7**: Results from splsda using mixOmics package accounting for repeated measures (repeated weeks 0, 2 and 8). **Panel A** is the loadings plot, where X and Y axes capture ~17% of total variance in X variables, and the plot shows distribution of high and low-fat burners (HB and LB). **Panel B** shows the cluster components (CC) on the Y axis and the variance that was captured in Loadings 1 and 2 on the X axis. Panel C is the VIP plot depicting the clusters that captured the most variance.

Supplemental Methods

*Plasma fatty acids analysis: Extraction and Derivatization.* After randomization, plasma (50 µL) was enriched with a rare or deuterated free fatty acid, a cholesteryl ester, a triglyceride, and a phospholipid and extracted with 10:8:11 cyclohexane: isopropanol: ammonium acetate[38,39]. Extracted lipids were either trans-esterified by sequential incubation in alkaline and acidic methanol, or directly methylated with trimethylsilyl-diazomethane in hexane in the presence of 10(*E*)-pentadecenoic acid to confirm derivatization efficiency[40]. Derivatized solutions were enriched with C23:0 as an internal standard and stored at -20°C until analysis.

FAMEs were separated on a 30m x 0.25mm, 0.25 µm DB-225ms in a 7890B gas chromatogram interfaced with a 5977B mass selective detector (Agilent Technologies, Santa Clara, CA) and quantified against 6–8-point calibration curves acquired and processed with MassHunter v. B 08[38]. Calibrants and internal standards were purchased from NuchekPrep (Elysian, MN), Sigma-Aldrich (St. Louis, MO), or Avanti Polar Lipids Inc.
